# Normalized LST is an efficient biomarker for homologous recombination deficiency and Olaparib response in ovarian carcinoma

**DOI:** 10.1101/2022.08.22.22278669

**Authors:** Yann Christinat, Liza Ho, Sophie Clément, Catherine Genestie, Jalid Sehouli, Antonio Gonzalez Martin, Ursula Denison, Keiichi Fujiwara, Ignace Vergote, Germana Tognon, Sakari Hietanen, Isabelle Ray-Coquard, Eric Pujade-Lauraine, Thomas A. McKee

**Affiliations:** Hôpitaux Universitaires de Genève, Department of Clinical Pathology, Geneva, Switzerland; Université de Genève, Geneva, Switzerland; Gustave Roussy, Paris, France; Charité - Universitätsmedizin Berlin (CVK), Berlin, Germany; MD Anderson Cancer Center Madrid, Madrid, Spain; Klinik Hietzing, Institute for gynaecological oncology und senology, Vienna, Austria; Saitama Medical University International Medical Center, Saitama, Japan; University Hospitals Leuven and Leuven Cancer Institute, Leuven, Belgium; Spedali Civili di Brescia, Brescia, Italy; Turku University Hospital, Department of Obstetrics and Gynecology, Turku, Finland; Centre Leon Bérard, Lyon, France; ARCAGY-GINECO, Paris, France

**Author notes:** **Corresponding Authors** Yann Christinat,; Thomas A McKee. **Fundings:** This study was partially funded by AstraZeneca, Cambridge, UK and Merck Sharp & Dohme LLC, a subsidiary of Merck & Co., Inc., Rahway, NJ, USA.

**Keywords:** HRD, PARP inhibitors, Ovarian cancer, Olaparib, Precision oncology, Myriad

## Abstract

**BACKGROUND:** The efficiency of the Myriad Homologous Recombination Deficiency (HRD) test to guide use of PARP inhibitors has been demonstrated in several phase III trials. However its high failure rate and limited accessibility establish a need for a clinically validated laboratory developed test.

**PATIENTS AND METHODS:** A novel biomarker to identify HRD was developed using TCGA data and, as part of the ENGOT HRD European Initiative, applied to 469 samples from the PAOLA-1/ENGOT-ov25 phase 3 trial using the OncoScan™ CNV Assay. Results were compared to the Myriad myChoice Genomic Instability Score (GIS) with respect to the progression-free survival in the Olaparib+Bevacizumab and placebo+Bevacizumab arms.

**RESULTS:** Analysis of the TCGA cohort revealed that a normalization of the number of large-scale state transitions (nLST) by the number of whole genome doubling events allows a better separation and classification of HRD samples than the GIS. The Oncoscan+nLST test yielded a lower failure rate on the 469 PAOLA-1 samples (10/469 vs 59/469 inconclusive results) and positive and negative agreement values of 96% (204/213) and, 81% (159/197) respectively. In nLST-positive samples, the hazard ratio (HR) was 0.40 (95% CI: 0.28-0.57) compared with 0.35 for Myriad GIS. In tumors that were BRCA wild-type and nLST-positive, the HR was 0.53 (Myriad: 0.41). In this subpopulation the nLST test and the Myriad myChoice test yielded a similar 1-year PFS (87% and 88%) but a different 2-year PFS (52% vs 60%) upon Olaparib+ Bevacizumab treatment.

**CONCLUSION:** The proposed test is a viable alternative to the Myriad myChoice HRD test and can be easily deployed in a clinical laboratory for routine practice. The performance is similar to the commercial test but its lower failure rate allows a 10% increase in the number of patients who will receive a conclusive laboratory result.

**Highlights:**

- A novel laboratory-developed test that increased HRD scoring accessibility for ovarian cancer patients
- Clinically validated on 469 samples from the PAOLA-1 trial within the ENGOT HRD European Initiative
- A similar performance as the Myriad myChoice Dx HRD test but at a lower failure rate

## Introduction

There is an unmet need for an academic assay to detect homologous recombination deficiency (HRD) in high grade serous ovarian, and perhaps in other cancers. The available approaches have shortcomings with respect to their technical characteristics and their cost. Ovarian Cancer is the eighth most common cancer affecting women worldwide^1^ and the majority present with advanced disease^2^.

Numerous clinical studies have shown that patients whose tumors have HRD, with or without germline or somatic breast cancer type 1 and 2 susceptibility protein mutations (gBRCA and sBRCA), have improved survival if treated with inhibitors of poly (ADP-ribose) polymerase (PARPi), particularly as maintenance therapy (reviewed by Foo *et al*.^3^). The rationale for this approach, known as synthetic lethality, is that BRCA mutations and PARPi target different DNA repair mechanisms and the loss of both is incompatible with cellular viability^3,4^.

Prediction of a response to PARPi is currently based on two strategies – detection of mutations in the genes involved in HRD (particularly *BRCA1/2*) and identification of a HRD phenotype. The advantage of mutation testing is that it provides a simple readout (presence or absence of a pathogenic or probably pathogenic mutation in the tested genes), though the presence of bi-allelic alterations is probably important. The weaknesses of this approach are that it depends on the completeness and accuracy of variant annotation databases and other mechanisms of gene silencing are overlooked. This was highlighted in the PAOLA-1 study where 19% of patients were considered to be BRCA wildtype but HR deficient and responded to Olaparib+Bevacizumab (more extensive mutation testing failed to reduce this percentage significantly)^5,6^. The advantage of identifying the HRD phenotype is that this approach detects the effect of diverse mechanisms of homologous recombination loss and is thus more sensitive as was confirmed in the PRIMA^7^, ARIEL3^8^, VELIA^9^ and PAOLA-1^5^ studies. Several techniques have been developed to identify HRD; the quantification of large scale genomic alterations (loss of heterozygosity (LOH)^10^, telomeric allelic imbalance (TAI)^11^ and large scale state transition (LST)^12^), specific mutational signatures^13^ and the absence of RAD51 foci^14,15^, certain of which alone or in combination are the basis of current assays. HRD is a frequent alteration identified in a wide range of tumors but particularly those with a genetic predisposition associated with gBRCA mutations.

Whole genome doubling (WGD) is another common genomic phenotype that is found in around 30% of advanced neoplasms. It is thought to promote tumorigenesis by facilitating genomic instability and helping to buffer against the negative effects of deleterious mutations. WGD has been shown to be particular prevalent in high grade serous ovarian cancers^16^. It is self-evident that the presence of a near doubling of genetic material in a tumor is a confounding factor for HRD analyses that depend on quantification of genomic alterations.

We report an academic laboratory developed, clinically validated test to detect HRD. The algorithm is publicly available within an R package on GitHub (https://github.com/yannchristinat/oncoscanR) to allow wide dissemination of this essential diagnostic modality.

## Methods

### TCGA cohorts

The TCGA Pan-cancer cohort (pancan12) contains 457 high-grade serous ovarian carcinomas (264 with an associated BRCA1/2 mutation status) and 112 triple negative breast carcinomas. CNV segments and the number of whole genome doubling events computed by the ABSOLUTE software were downloaded from https://www.synapse.org/#!Synapse:syn171046417. Mutational status for *BRCA1* and *BRCA2* were obtained from the TCGA article^18^. ER, PR, and Her2 status for the breast cancer samples were downloaded from https://www.cBioPortal.org on May 18, 2020.

### PAOLA-1 cohort patient samples

The PAOLA-1 study is part of the European ENGOT (European Network of Gynecological Oncology Trial) HRD initiative led by the French group GINECO (Groupe des Investigateurs Nationaux pour l’Etude des Cancers de l’Ovaire)^19^. To allow the 10 initial participant laboratories to analyze the same samples from the PAOLA-1 trial^5^, samples with the highest content of DNA were prioritized (ARCAGY-GINECO tumor bank, Institut Curie, Paris). DNA was extracted from FFPE tumor slides, preferentially from patients untreated before surgery. 100 ng tumor DNA samples from each patient were transferred to 96-well plates that were sent to Geneva at -80°C, after agreement from the French authority. BRCA wild type tumors were selected for the first phase of 85 samples that was designed to test the correlation with the MYRIAD test, specifically for those BRCA wild type tumors where the GIS score has been shown to detect HRD positive cases. The second phase involved testing 384 additional samples with high DNA content.

Care was taken that this selection was representative of the global PAOLA-1 population in terms of BRCA and HRD status distribution and that the benefit of Olaparib + bevacizumab maintenance versus bevacizumab among the 469 selected cases was in the same range as that of the global PAOLA-1 population of 806 cases. These 469 samples (female patients with a high-grade ovarian adenocarcinoma; median age: 60) were analyzed with the Oncoscan FFPE Assay Kit (cat. 902695; ThermoFisher Scientific) following the manufacturer’s instructions. The arrays were stained in GeneChip Fluidics Station (ThermoFisher Scientific) and scanned using the Gene Chip scanner (ThermoFisher Scientific, Waltham, Massachusetts, United States). CEL files (Affymetrix DNA microarray image analysis software output) generated from the scanned array image were converted to Oncoscan array data (OSCHP files) and analyzed using Chromosome Analysis Suite (ChAS) software (version 4.0; ThermoFisher) with reference files NetAffxGenomicAnnotations.Homo_sapiens, hg19; NA33.r1. The copy number variation segments were evaluated manually in ChAS and exported in a text file format. Parameters in ChAS were adapted from the default settings to exclude segments containing less than 50 markers or smaller than 50 kbp (kilo base pairs).

### HRD scores computation

The number of Large-scale State Transitions (LST) is computed as described in the paper from Popova *et al*. [Popova, Can. Res. 2012]. First, segments smaller than 3Mb are removed, then segments are smoothed with respect to copy number at a distance of 3Mb. The number of LSTs is the number of breakpoints that have a segment larger or equal to 10Mb on each side.

The LOH score is computed as described by Abkevich *et al*.^10^: i.e. the number of segments larger than 15Mb with loss of heterozygosity but excluding segments spanning an entire chromosomal arm (computed as previously described^20^).

The TAI score is computed according to Birbak *et al*.^11^: the number of telomeric segments that display an imbalance between the maternal and paternal alleles and that are larger than 11Mb but do not cover more than 90% of the chromosomal arm.

Three normalization methods for the LST were investigated.

- A normalization by the average copy number 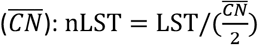
- A normalization by the ploidy as proposed by Timms *et al*.^21^: nLST = LST − kP where P is the ploidy of the sample, estimated as the median copy number across the genome, and k is a factor.
- A normalization by the number of whole genome doubling events (WGD): nLST = LST − kW. The number of WGD, *W*, is estimated as proposed by Carter *et al*.^17^: W=0 if the 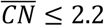, W=1 if 2.2 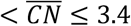 and 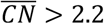

Of note, copy number-neutral segments with a loss of heterozygosity (LOH) are excluded from the nLST computations to improve the sensitivity in samples with low tumor content. A sample was suspected of having low tumor content whenever there was less than 2 LST and no arm-level alterations.

All statistical analyses and graphs were performed with R 3.5.1^22^ and the packages ‘forestplot’, ‘survival’ and ‘survminer’. The WGD-based nLST score and other Oncoscan-related analyses were performed with the oncoscanR v1.0.0 package (https://github.com/yannchristinat/oncoscanR).

## Results

### The number of LST depends of the number of WGD events

Using the public datasets from the TCGA (457 ovarian carcinoma and 112 triple negative breast cancer patients), we observe that the distributions of the three components of the HRD score from Telli *et al*. (as used in the Myriad myChoice test)^23^ shift with the number of whole-genome doubling events (WGD) (Figure 1). The number of LSTs and the number of TAIs increase with the number of WGD events whereas the number of loss of heterozygosity events (LOH) diminishes. Interestingly, the sum of the three markers does not seem to be affected by the number of WGD events as the peaks of the bimodal distributions are aligned. However the percentage of HRD-positive cases (defined as LST+LOH+TAI≥42) decreases as the number of WGD events increases: 40% (102/255), 23% (57/245) and 13% (9/69) for respectively no WGD, one WGD and two WGD events. Therefore, a test based solely on the presence or absence of WGD events will have a positive predictive value of 40% (102/255) and a negative predictive value of 79% (248/314) where we would expect 30% and 70% by chance (Cohen’s Kappa of 0.196).

**Figure 1.**
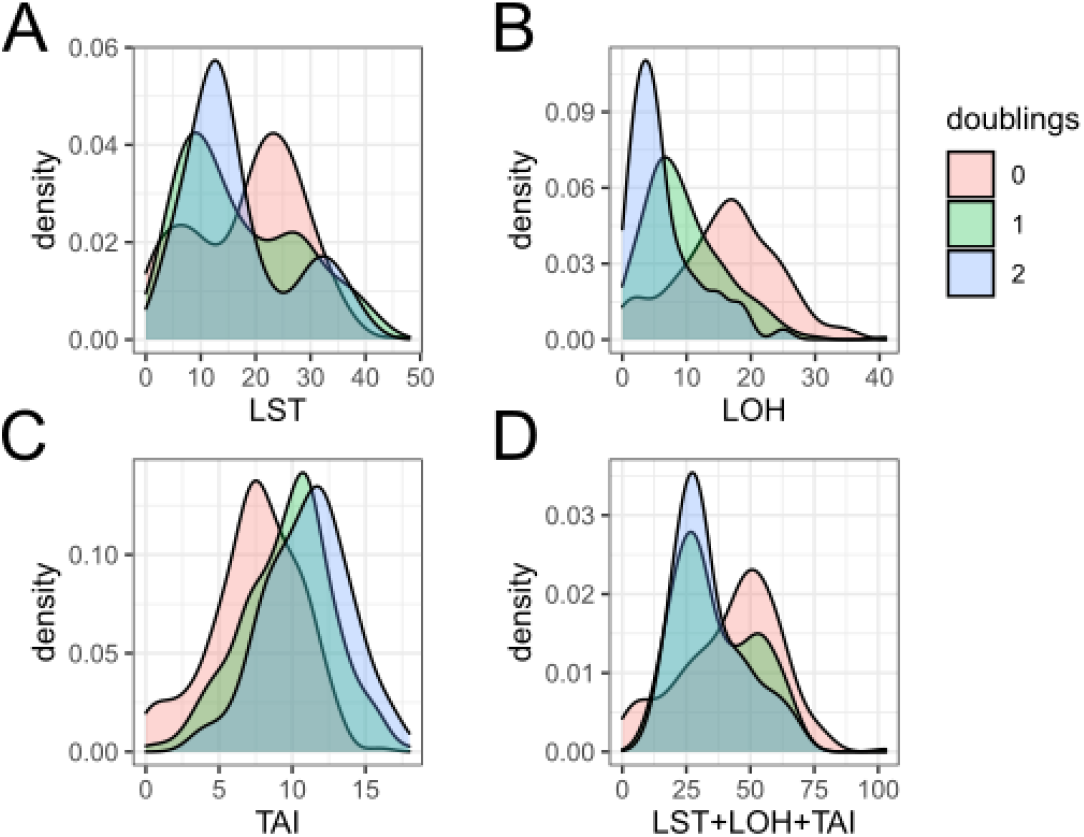
Distribution of A) LST, B) TAI, C) LOH scores and D) their sum on the TCGA cohort (OV+TNBC), with respect to the number of whole genome doubling events as given by the ABSOLUTE software.

### The number of LST normalized by the number of WGD events is an efficient biomarker for HR-deficient tumors

Since the number of LST is the only value to have a bimodal distribution, as expected for a biomarker identifying two distinct populations, we postulated that the number of LOH and TAI events in the HRD score from Telli *et al*.^23^ act as surrogates for a normalization by the ploidy or the number of WGD events but do not add value to a normalized LST score. To test this hypothesis, we investigated three normalization methods for the LSTs:

- A ploidy-based method, as suggested by Timms *et al*.^21^ that shifts the LST values in function of the ploidy of the sample.
- A method based on the average copy number that, instead of shifting the values by a discrete number, normalizes the LST directly by the amount of DNA in the cell.
- A WGD-based method, similar to the Timms *et al*.’s, but that uses the estimated number of WGD events (none, one, or two) instead of the ploidy.

As the HRD-negative LST peak of the no-WGD-events samples has a value of 7 (Figure 1A), it was chosen as the baseline parameter for the ploidy- and WGD-based normalization methods (k=7/2). Of note, the suggested k=15.5 for the ploidy-based normalization method shifts all polyploid samples into the first peak and was not further considered (Suppl. Figure S1).

These methods were compared using two performance metrics:

- The BRCA detection rate, i.e. the percentage of samples with a homozygous mutation on *BRCA1* or *BRCA2* within the positive cluster. A good test is expected to classify all BRCA-mutated samples as positive. (Of note, this measure was only computed using the 264 ovarian case that had been characterized for BRCA mutations.)
- The between clusters sum of squares (BCSS), which measures the average distance between two clusters. A good test is expected to have a clear separation and thus a large BCSS.

Figure 2A shows that a normalization by the number of WGD events yields better results than a normalization by the ploidy or the average copy number, independent of the test cutoff. The number of normalized LST, irrespective of the normalization method, also allows a better classification of BRCA-mutated samples and a better cluster separation than the HRD score from Telli *et al*.

**Figure 2.**
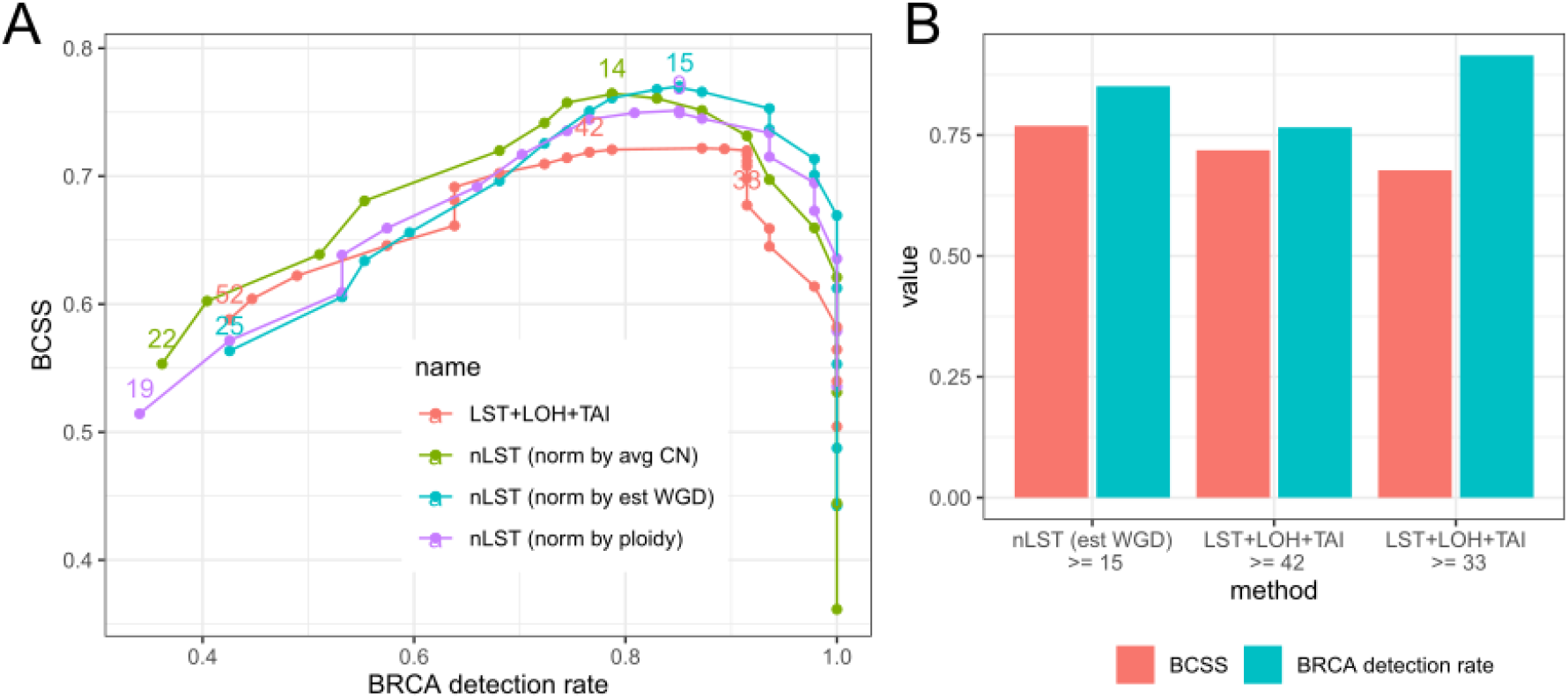
Evaluation of different methods on the TCGA cohort (OV+TNBC). A) Results at different cutoffs. B) Results at the recommended cutoffs.

For the number of LST normalized by the WGD events, a cutoff point at 15 yields the highest BCSS and was chosen for the test. Figure 2B, shows that the nLST method using the cutoff of 15 results in a better BRCA detection and cluster separation than the Myriad-like test at their recommended cutoff of 42.

### The Oncoscan+nLST test yields a much lower failure rate than Myriad on PAOLA-1 samples

The percentage of inconclusive tests stands at 13% (59/469) for the Myriad GIS score (reduced to 9% when the BRCA mutational status is included) while the Oncoscan assay yielded a failure rate of 2% (10/469). Among the 59 Myriad GIS inconclusive samples, 5 samples also failed with the Oncoscan assay (probably due to DNA quality issues), 21 were suspected to have an extremely low tumor content (<15%) and 13 were estimated to have a low tumor content (15-20%). Overall, 27 samples were predicted HR-proficient (negative) by the Oncoscan+nLST test but were likely inadequate in terms of tumor content. An analysis of the nLST value distribution in samples with low tumor content revealed a distribution bias as the tumoral content reaches 15% (T-test p-value of 1.69e-5 for 15% vs >20% and 0.204 for 20% vs >20%; Suppl. Figure S2). However, a bimodal distribution was still observed from 15% to 20% and all positive samples were also predicted positive by Myriad. The limit of detection thus stands at 20% tumor content but a positive call at 15% tumor content can be trusted.

### The nLST score is highly correlated with the Myriad GIS

On the 405 patients from the PAOLA-1 trial with a conclusive result from both tests, we observe a tight correlation between the nLST score and the Myriad instability score (Pearson’s correlation coefficient of 0.88, p-value < 2.2×10^−16^; Suppl. Figure S3).

With the default threshold of 42 for the Myriad GIS, the nLST score (positive if ≥15) yielded a Cohen’s Kappa of 0.79 and positive and negative agreement values of 96% (204/211) and 81% (159/195) respectively (Table 1). However, with the threshold set at 33 for the Myriad score the Cohen’s Kappa increases to 0.83, indicating an almost perfect agreement.

**Table 1.** HRD calls comparison (nLST vs Myriad GIS) on 469 patients from the PAOLA-1 trial. Low %T denotes the number of samples labeled HRD-negative by the nLST test but where the tumoral cellularity was likely too low for the Oncoscan analysis.

|  |  | *nLST*≥15 |  |  |
| --- | --- | --- | --- | --- |
|  |  | *Positive* | *Negative (low %T)* | *Inconclusive* |
| *GIS*≥42 | *Positive* | 204 | 6 (1) | 3 |
|  | *Negative* | 36 | 159 (5) | 2 |
|  | *Inconclusive* | 12 | 42 (21) | 5 |

The nLST test is thus an excellent surrogate for the Myriad test, with a higher similarity with the 33 threshold, but with a much lower failure rate.

### The nLST test is predictive of the response to Olaparib+Bevacizumab

The Myriad myChoice test includes a genomic instability score and the BRCA mutation status. The Oncoscan+nLST test, in contrast, is not designed to identify BRCA mutations but in practice the BRCA mutational status is already known or is assessed via sequencing. The PFS hazard ratio with respect to the treatment arm (Olaparib+Bevacizumab or Placebo+Bevacizumab) and the test status (HRD-positive or negative) was compared in relation to the BRCA mutation status provided by the Myriad test (Figure 3). In the 151 BRCA-mutated samples, the hazard ratio was 0.36 (95% CI: 0.22-0.57). In the BRCA wild-type samples, the nLST test classified 113 samples as HRD-positive, which yielded a hazard ratio of 0.53 (95% CI: 0.33-0.86). In the same subpopulation, the Myriad GIS with a threshold at 42 and 33 produced a hazard ratio of 0.41 (95% CI: 0.24-0.70; 91 HRD-positive samples) and 0.49 (95% CI: 0.31-0.78; 120 HRD-positive samples) respectively. Irrespective of the BRCA status, the Myriad test with a threshold at 33 and the nLST test all yield very similar hazard ratios on the PFS when Olaparib is added to the Bevacizumab maintenance treatment. Interestingly, the addition of the BRCA status to the nLST score or the Myriad GIS does not improve the hazard ratio (from 0.40, 95% CI: 0.28-0.57, to 0.43 for the nLST). Three BRCA-mutated nLST-negative patients received Olaparib and all had a low Myriad GIS and a low PFS (7.9, 13.9 and 18.7 months). In particular, the patient with the lowest PFS had a heterozygous mutation which concurs with the negative HRD evaluations in indicating that there may be no deficiency in the HR pathway.

**Figure 3.**
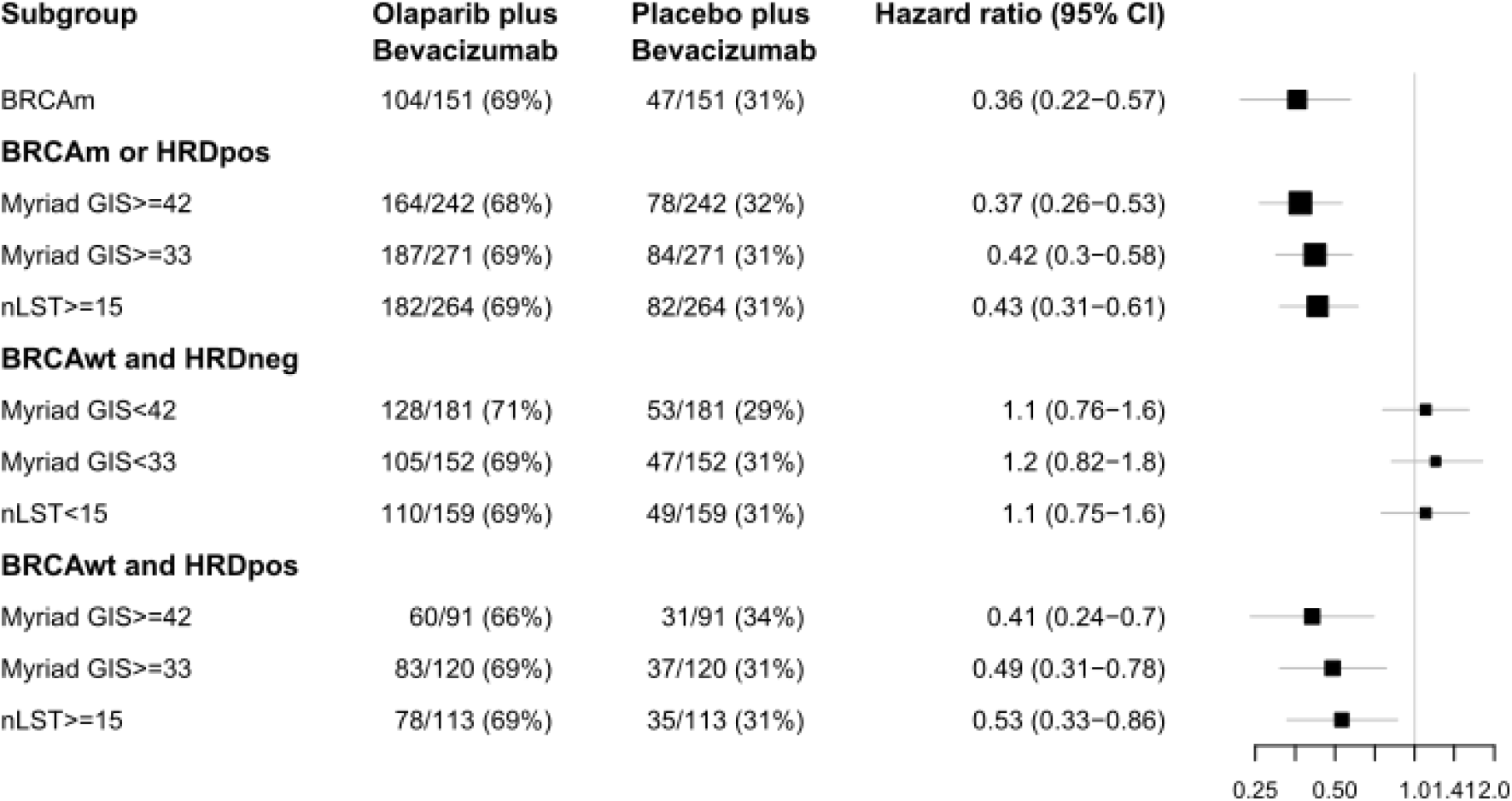
Hazard ratio for HR-proficient (HRD-negative) and HR-deficient (HRD-positive) samples, according to different tests on 405 PAOLA-1 patients with conclusive results from all tests, stratified by the BRCA mutation status as given by Myriad.

### BRCA wild-type patients with an intermediate nLST score derive a short-term benefit from Olaparib+Bevacizumab

The nLST test yielded more HRD-positive samples in the BRCA wild-type population than the Myriad test with a threshold at 42. These nLST-positive but Myriad GIS-negative samples display an intermediate PFS between the positive and negative samples (hazard ratio: 0.71, 95% CI: 0.26-1.90; 28 samples). The PFS difference arises almost entirely in the second year after the start of treatment. In this BRCA wild-type population treated with Olaparib+Bevacizumab the nLST test and the Myriad test yielded a similar 1-year PFS (87% and 88%) but a different 2-year PFS (52% vs 60%). As most nLST-positive but GIS-negative samples have a nLST score below 20 (third quartile at 19). We investigated a three-way classification: nLST<15 (negative), 15≤nLST<20 (positive/mid), nLST≥20 (positive/high). The negative samples yielded a 1-year PFS of 67% (95% CI: 59-76%). The positive/mid samples yielded a 1-year PFS of 85% (95% CI: 73-100%), which is close to the 1-year PFS of the positive/high samples: 89% (95% CI: 81-98%). By contrast, the 2-year PFS of the positive/mid samples is closer to the negative samples than the positive/high ones (0.30 [95% CI: 17-53%], 24% [95% CI: 17-33%], and 66% [95% CI: 54-80%] respectively). Figure 4 shows that this nLST-intermediate population is enriched in patients with tumor recurrence during their second year of treatment. This behavior was not observed with a three-way classification of the Myriad GIS (positive/mid if 33≤GIS<42; Suppl. Figure S4).

**Figure 4.**
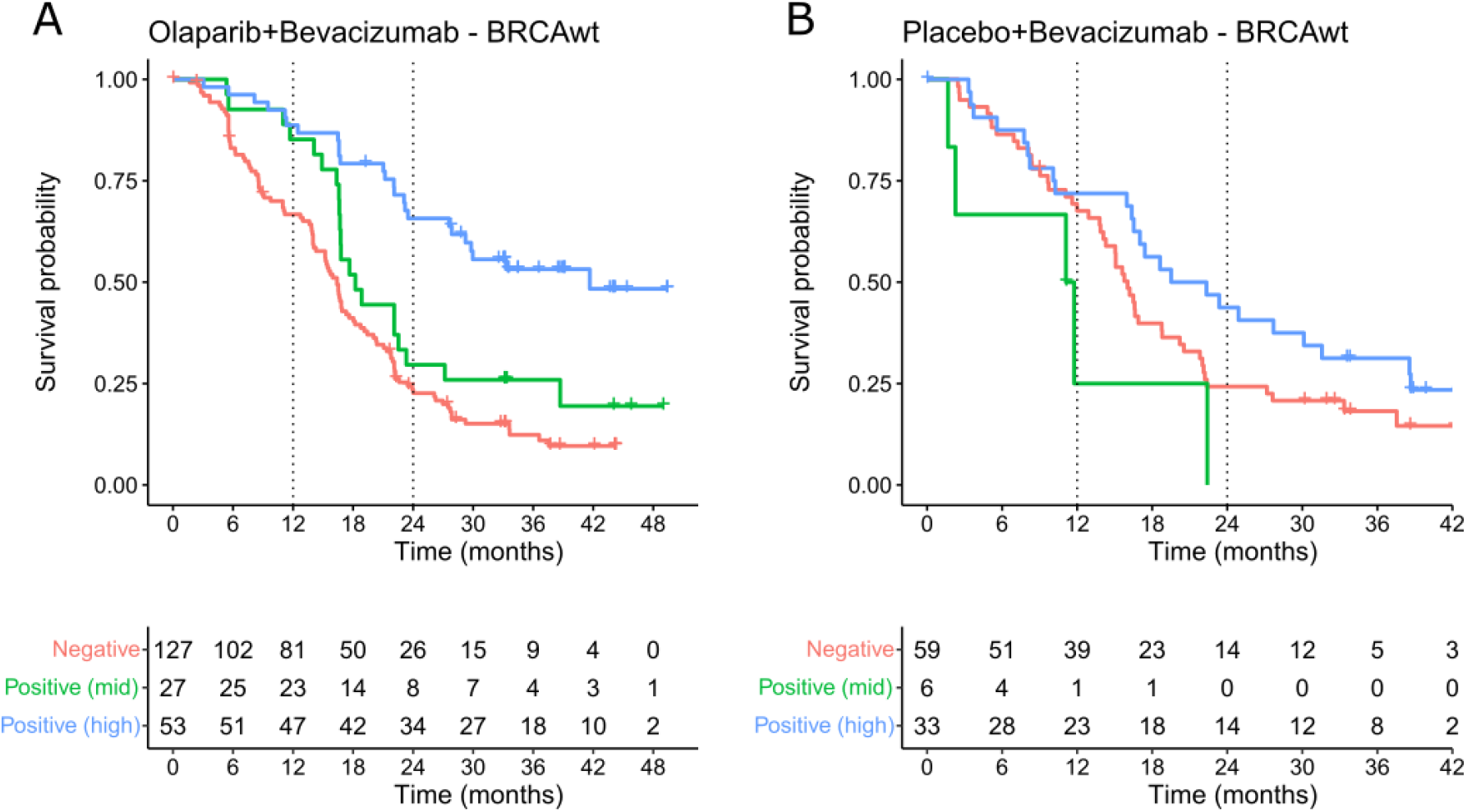
Kaplan-Meier plot of a three-way classification of the nLST score, with thresholds at 15 and 21, in the BRCA wild-type population upon A) Olaparib+Bevacizumab and B) Placebo+Bevacizumab treatments.

## Discussion

Many studies have shown that inhibition of PARP is an effective treatment that prolongs OS and PFS for high grade serous carcinoma of the ovary^3^. The efficacy of this treatment is in great part dependent on the loss of the homologous recombination pathway for DNA repair^3,4^. However the adverse effects of PARPi are similar to chemotherapy and thus careful selection of patients is important^24^. Currently available assays are in the commercial sector leading to restricted access both for clinical and research applications. We report here the development and technical and clinical validation of a novel assay for HRD which is available to the academic community. This test has been integrated into our routine laboratory workflow and represents a low cost alternative to commercial assays. The algorithm on which it is based could be applied to other data sources as long as confident calling of copy number can be assured. The absence of copy-neutral LOH testing in the nLST score also makes it a promising candidate for other technology platforms but the concordance with the Oncoscan platform would have to be confirmed. The Oncoscan technology is particularly well adapted to calculating the HRD value using FFPE tissues and circumvents one of the main barriers to genomic testing, DNA quality. Indeed the microarray technology results in no enzymatic reactions being performed on the tumor DNA^25^, in contrast to the capture-based Next-Generation Sequencing assays probably in part explaining the increase in the number of conclusive results. Low tumor cell percentage, remains a barrier to accurate genomic analysis and we demonstrated that our assay requires between 15% and 20% of tumor cells in the sample in order to give a result with confidence.

We find that calculating LST is an efficient method to identify the genomic scars induced by the loss of HR and that normalizing the LST using genomic doubling events predicts response to Olaparib therapy, in addition to Bevacizumab, as well as the Myriad myChoice test in 469 samples from the PAOLA-1 study, classifying more patients as HRD-positive than the Myriad test. However one cannot exclude that this performance is due to the combination of Olaparib and Bevacizumab as the Myriad test did not perform as well in PARPi-monotherapy trials. The set of patients that were positive for the nLST score and negative for the Myriad myChoice test represent an interesting subgroup having an intermediate nLST score (between 15 and 20 nLST). They displayed a better response to Olaparib+Bevacizumab than patients that were negative for both tests, but a worse long term response than the patients predicted HRD-positive by both tests. This could represent a mixture of responders and non-responders that were not efficiently separated by the test however the data suggest that this set of patients display an initial response to PARP inhibitors that is not sustained, suggesting both a biological basis for this observation and the existence of partial responders despite a low number of patients in this group. Indeed, it raises the question of whether a 3-class system has a clinical utility, a suggestion supported by the existence of hypomorphic BRCA mutations^26^ and the fact that different thresholds for the Myriad test have been used in clinical studies^27^. The inclusion of these patients in the HRD-positive category would have a negative effect on the global hazard ratio of the test, as observed in our case, but it still translates into a 12-month benefit for patients that receive treatment.

The addition of the BRCA mutation status to the nLST score or the Myriad GIS score, as performed in the Myriad myChoice test, did not improve the performance of these tests. In ovarian cancer, more than 90% of the BRCA-mutated tumors show a genetic alteration on the second allele and mutations are thus homozygous. This leaves a small group of patients that have a BRCA mutation possibly without a HR deficiency. In the PAOLA-1 sub-cohort, we observed two cases with a heterozygous mutation and low GIS and nLST scores that did not respond well to the Olaparib treatment. The BRCA status, which is in general derived in parallel with the HRD score, is however beneficial in samples with a low tumor cell content as it eliminates some false negatives. It might also remove some false positives if a reversion mutation is detected. In our opinion, the BRCA status should be considered as a complement to HRD testing and the zygosity of the mutation should be taken into account in cases with a low HRD score.

We have developed and make available an academic assay for HRD detection that complements available assays, and identifies a new group of ovarian cancer patients who seem to respond to PARP inhibition but relapse earlier. We expect this assay to facilitate the availability of HRD testing and thus improve patient wellbeing.

## Supporting information

REMARK checklist

## Data Availability

All data in the present study produced from the TCGA cohort are available upon reasonable request to the authors.
All data in the present study produced from the PAOLA-1 trial data are unavailable.

## Acknowledgment

We would like to thank AstraZeneca UK and Merck Sharp & Dohme for their financial support, ARCAGY for having put in place this wonderful HRD initiative and having invited us to participate in it, and the whole molecular pathology laboratory in Geneva for their quality work with the Oncoscan assay.

