## Supplementary Figures for "Normalized LST is an efficient biomarker for homologous recombination deficiency and Olaparib response in ovarian carcinoma"

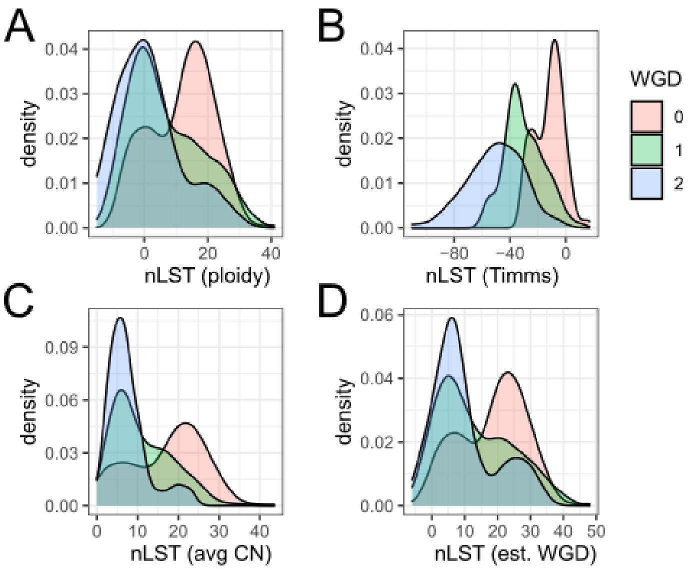


***Figure S1***. *Normalized LST score distribution according to different methods and the number of WGD events as given by the ABSOLUTE software. A) Ploidy-based method with k=3.5. B) Ploidy-based method with k=15.5. C) Method based on the average copy number. D) WGD-based method.*

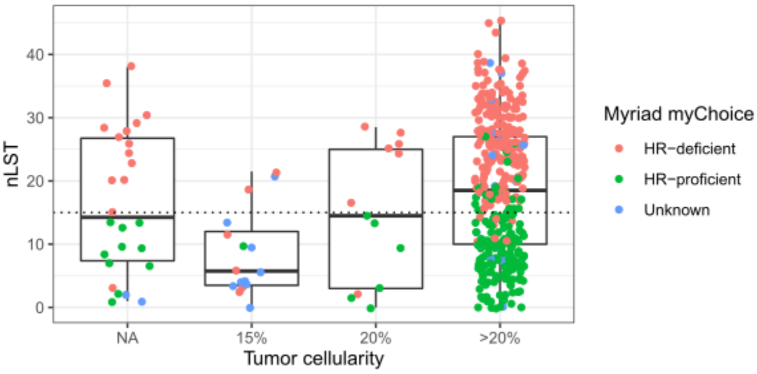


***Figure S2.*** *Distribution of nLST scores with respect to the tumor content as estimated by the Oncoscan analysis software.*

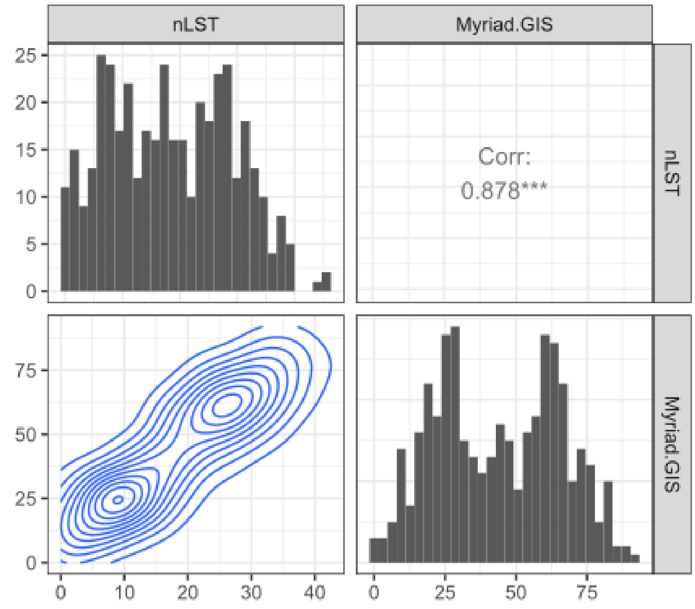


***Figure S3.*** *Distribution for the nLST score and Myriad GIS on 405 patients from the PAOLA-1 trial and their paired comparison.*

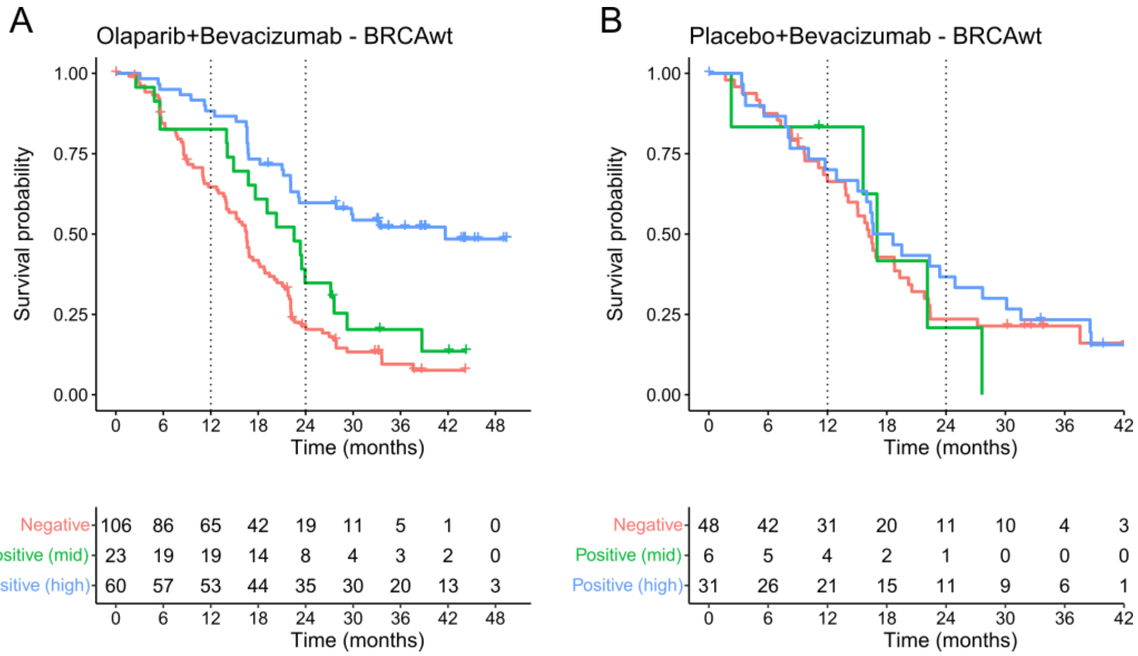


***Figure S4.*** *Kaplan-Meier plot of a three-way classification of the Myriad GIS score, with thresholds at 33 and 42, in the BRCA wild-type population upon A) Olaparib+Bevacizumab and B) Placebo+Bevacizumab treatment.*
